# Knowledge, perceptions and acceptance of COVID-19 Vaccine in Plateau States, Nigeria: A Qualitative Study

**DOI:** 10.1101/2025.10.16.25338166

**Authors:** Sophia Osawe, Sussan Israel-Isah, Timothy Adejoh, Tunde A. Alabi, Felicia Okolo, Sikiratu Babamale, Adam Abdullahi, Alash’le Abimiku

**Affiliations:** International Research Centre of Excellence, Institute of Human Virology Nigeria (IHVN) Nigeria; Department of Sociology, University of Lagos, Nigeria; Plateau State Human Virology Research Centre, Jos, Nigeria; Institute of Huma Virology, School of Medicine, University of Maryland, Baltimore, USA

**Keywords:** Vaccine hesitancy, Vaccine acceptance, Pregnant women, Healthcare workers, Policymakers

## Abstract

**Introduction and Objectives:** Knowledge, perceptions, and acceptance of vaccines play a role in evaluating the progress of past immunization programs and planning for future pandemics. The study assessed the knowledge, perceptions, and acceptance of COVID-19 vaccines among pregnant women, HCWs, and policymakers in Nigeria and identified the barriers and enablers to the uptake of COVID-19 vaccines and future ones.

**Methods:** The study adopted qualitative methods of qualitative collection, comprising focus group discussion (FGD) with pregnant women-who were HIV positive and negative- and key informant interviews (KII) with healthcare workers (HCWs) and policymakers in Plateau State, Nigeria. The sample comprised forty pregnant women, five HCWs and five policymakers. Data was managed using Dedoose (version 9.0) and analyzed thematically.

**Results:** Although all sub-groups acknowledged the importance of vaccination, COVID-19 vaccine hesitancy was identified amongst a significant proportion of pregnant women. This was because of safety concerns, misinformation and fear of side effects. HCWs were identified as key influencers of vaccine acceptance during pregnancy. Also, family and friends, personal beliefs, and awareness were found to be influencers of vaccination, while religious and cultural beliefs were identified as barriers. Policymakers and HCWs believe that there is a need for education and awareness to address misinformation and improve the trust in vaccine safety and the health system among pregnant women.

**Conclusion:** The participants were knowledgeable of vaccines. However, pregnant women expressed skepticism about the safety of the COVID-19 vaccines. Thus, addressing misinformation through public health awareness programs is important while leveraging the influence of the health workforce, community, and religious leaders in promoting vaccine acceptance.

## BACKGROUND

The COVID-19 pandemic tremendously impacted public health, disrupting health systems and health service delivery to vulnerable populations, including pregnant women in Nigeria. Pregnant women were particularly vulnerable to COVID-19 infections due to the dynamic immunological and physiological changes that occur during pregnancy (1). This risk is further heightened in individuals with HIV, as the infection can compromise immune function, making them more susceptible to severe disease outcomes (2). The rapid development and deployment of COVID-19 vaccines were important public health strategies adopted to curb the risk of infection and poor health outcomes especially amongst vulnerable populations. However, vaccine hesitancy amongst pregnant women is a major public health concern, especially in low-and-middle-income-countries (LMIC) like Nigeria (3–5). Although Nigeria achieved 70% coverage for COVID-19 vaccination, it is important to understand the knowledge, perceptions and acceptance of vulnerable populations towards COVID-19 vaccines (6).

The World Health Organization (WHO) emphasizes the importance of addressing misinformation to enhance vaccine uptake (7). Generally, HCWs play very vital roles in influencing vaccine uptake, as they act as sources of information to pregnant women. As a result, poor knowledge and attitude of HCWs towards vaccines can influence misinformation. The impact of misinformation on vaccine hesitancy was profound, with many individuals relying on social media rather than the healthcare system for health information due to its easy access through this platform during the early period of the pandemic (8). Studies have shown that poor awareness, cultural beliefs, misinformation, concerns about vaccine safety, and the influence of HCWs shape perceptions, acceptance, and uptake of the COVID-19 vaccine among the general population and pregnant women (7–11). A study found that misinformation about vaccine safety and efficacy contributes to hesitancy, particularly among pregnant women (9)Thus, adequate awareness and positive perceptions about COVID-19 may motivate HCWs to share vital information that will promote vaccine acceptance.

In many regions of the world, the rapid development of COVID-19 vaccines led to increased skepticism regarding the safety and efficacy of the vaccines (Rosenthal & Cummings, 2021). A multinational study indicated that conspiracy beliefs significantly affected vaccine acceptance across various countries, including Asia and Africa (10). Similarly, a scoping review highlighted that barriers to COVID-19 vaccine uptake include socio-economic factors, cultural beliefs, and limited access to healthcare services (11). The role of policymakers in decision-making and gaining the public’s trust in accepting vaccines is crucial to ensuring vaccine acceptance. Thus, assessing an interplay between the knowledge and perceptions of pregnant women, policymakers, and HCWs can provide valuable insights for identifying the barriers and enablers to the uptake of COVID-19 vaccines and managing future pandemics.

This study investigated the knowledge, perceptions, and acceptance of COVID-19 vaccines amongst pregnant women, HCWs, and policymakers in Nigeria. This study provides insight into the roles of critical stakeholders in influencing vaccine uptake amongst vulnerable populations such as pregnant women. While the pandemic might be over, this study can inform public health strategies and policies to improve maternal immunization programs for subsequent infectious disease outbreaks. This study’s findings can help promote vaccine acceptance and prevent the negative impact of subsequent infectious diseases on maternal and fetal health.

## METHODS

### Study Design and Population

The study adopted a qualitative method, which comprised FGDs and KIIs. The study location was Plateau State, North-Central region of Nigeria and data were collected from June to July 2024. The study was part of a broader research project that assessed COVID-19 vaccine acceptance among Nigerian women in one clinical site in Plateau State. The study was funded by the National Institute of Health (NIH).

The study population comprised pregnant women, HCWs (doctors, nurses) and policymakers. For the FGD, only pregnant women aged 16 or older and receiving antenatal care in the selected health facilities were eligible to participate. For the KIIs, HCWs who were 18+ years of age and were working in the selected health facility were eligible to participate in the study. Health policymakers who resided in the states and were familiar with the study location participated in the study.

### Sampling

A purposive sample was used to select antenatal clinic in PLASVIREC hospital for the study. A total of four FGDs were conducted with pregnant women, and each one comprised 10 women, making a total of 40. Two FGD groups comprised women living with HIV and the other two without HIV. Five HCWs and Five policymakers were purposively selected for the KIIs. Potential participants were recruited from the selected health facility until the desired sample size was attained.

### Data collection

FGDs were conducted on pregnant women visiting the health facilities for ante-natal care, and KIIs were conducted with HCWs and policymakers. Trained qualitative interviewers conducted the interviews using piloted KII and FGD guides. Each qualitative interviewer underwent comprehensive training on the study methods, qualitative research techniques, and ethical standards. For the FGDs, the guide included questions on the level of awareness and uptake of vaccines in pregnancy, perceptions and misconceptions about COVID-19 vaccines, factors that contribute to decision-making to take vaccines, sources of health information, and recommendations to improve vaccination programs.

The KII guide for HCWs contained questions about recommended vaccines and uptake during pregnancy, perceptions about COVID-19 vaccines, the influence of HCWs’ motivation on pregnant women’s decision to take vaccines, perceptions about the impact of getting health information for COVID-19 from other sources, perceived factors that influence the decision to take vaccines and recommendation to prevent vaccine hesitancy. For the policymakers, the questions revolved around the recommended vaccines for pregnant women, the role of HCWs in influencing vaccine acceptance in pregnancy, perceptions about the influence of close relatives on pregnant women’s decision to take vaccines, perceived influence of health information sources and cultural factors on decision making and recommendation to improve vaccine hesitancy. All FGDs and interviews were held at PLASVEREC in a private location to ensure confidentiality and participants’ comfort. The interviews were audio-recorded and conducted both in Hausa and English; the KIIs lasted about 25 to 30 minutes in length and 60 to 90 minutes for the FGDs. The language of the interview was both English and Hausa.

### Data analysis

Deidentified audio recordings were translated and transcribed verbatim from Hausa into English in Microsoft Office Word. The data were managed with Dedoose (version 9.0). Mixed coding approaches were utilized to identify patterns, themes, and insights from the participants’ narratives about pregnant women’s decision-making in taking a vaccine for themselves and their infants. Using the deductive approach, two research team members developed a qualitative codebook based on the pre-existing study objectives and structural codes derived from the interview guides. An inductive approach was used to identify salient points in the data not captured in the code book. Regular meetings were held to resolve differences in the coding patterns. Findings were synthesized thematically.

### Ethical Considerations

An ethical approval to conduct the study was granted by National Health Research Ethics Committee (approval number NHREC/09/23/2010b). Potential participants were verbally informed to participate in the study at the antenatal clinics in the selected hospital. Eligible participants signed an informed consent form and were enrolled into the study. The anonymity and confidentiality of participants was ensured as no personal identifying information was collected. Participants were informed of their rights to withdraw from the study at any point they felt uncomfortable proceeding with the study. Prior to the interview started, participants were reminded again that the discussion would be audio recorded and any information they provided would be treated anonymously.

## RESULTS

The majority of the participants were between 18 and 35 years old. All of the women were married, and most of them were engaged in trade, followed by civil service employment. Nearly one-third had attained a secondary level of education. The predominant religious affiliation was Christianity, with a smaller proportion identifying as Muslim.

### Knowledge and Perception of Vaccines and COVID-19 Vaccination among Pregnant Women

The majority of the pregnant women demonstrated some knowledge of vaccines. The women knew about the tetanus vaccine and indicated receiving it during pregnancy. According to one participant, *“the Tetanus vaccine is to protect me and the child from tetanus”* (Pregnant woman, FGD). Although, pregnant women were aware of COVID-19 vaccination, their perceptions about the vaccines were fraught with skepticism and fear, with many expressing concerns about the safety of the vaccine during pregnancy. Several participants shared concerns related to adverse reactions and the negative side effects. In this regard, a participant believed*, “if someone takes the vaccine, it has an effect on the person”* (Pregnant woman, FGD). Another participant stated that, *“I didn’t want to take it because of the pregnancy” (Pregnant woman, FGD).* Another participant remarked thus: *“I thought maybe it will affect me and the unborn baby”,* suggesting the fear of losing the pregnancy if they took the vaccine. The belief that the COVID-19 infection was a hoax also contributed to hesitancy to COVID-19 vaccination. In this regard, a pregnant woman participant said that “I *personally did not really believe COVID-19 exists”* (Pregnant woman, FGD).

While most pregnant women accepted vaccines (anit-tetanus) administered by HCWs during ANC visits due to information, guidance and trust in the advice from HCWs,, very few took the COVID-19 vaccine during pregnancy, as majority cited worries about its impact on their health and that of their unborn children. participant

### Knowledge and Perception of Vaccines and COVID-19 Vaccination among Healthcare Workers

HCWs were knowledgeable and reported mixed perceptions about vaccines, while recognizing the importance of vaccination for pregnant women. Majority of the HCWs were of the opinion that pregnant women usually accept all vaccinations given to them during their ANC visits and throughout their pregnancy period, citing that they do this because they believe and accepts all recommended medication and treatment from the HCWs especially when they are informed of the benefits to their health and that of their infants. One HCW remarked, *“Our women are very cooperative usually everything you write for their antenatal clinic they accept, they accept readily” (HCW, KII)*.

HCWs were generally aware of COVID-19 vaccinations but expressed mixed feelings about their efficacy, acceptance, and uptake. While some supported COVID-19 vaccination for pregnant women, others were concerned about its safety, citing that the vaccine’s rapid introduction raises doubts about its efficacy and resulting in expressed fears of the long-term effects. A participant mentioned that *“we as a health worker, we explain to them the importance of taking the vaccine and most of them would want to know about the effect on their babies. I think that’s the major thing on their mind but when we make them to understand that is not going to affect their babies negatively, they will opt for it”*. Another participant stated that *“Some people think it is still at the experimental stage” (HCW, KII).* HCWs reported that misinformation from family and friends shape patients’ decisions regarding acceptance and uptake of COVID-19 vaccine. As one HCW mentioned that *“some say after taking the vaccine, they will die in two years”* (HCW, KII). HCWs believe that education and community engagement are vital in addressing vaccine hesitancy, advocating for continuous sensitization efforts to improve acceptance among pregnant women and those living with HIV.

### Knowledge and Perception of Vaccines and COVID-19 Vaccination among Policy Makers

According to policymakers, vaccinations that are safe and effective should be given to pregnant women, to prevent the onset of diseases for both mother and infant. They also believed that the COVID-19 vaccine is crucial for preventing infection and related morbidities among pregnant women. One policymaker stated that, *“I believe vaccines, as long as they are beneficial and safe in pregnancy, I think they should be recommended to the pregnant women so that they can also benefit”* (Policymaker, KII). However, policymakers raised concerns about the rapid roll-out of COVID-19 vaccines, with fears about the long-term effects and misinformation influencing public perception, thus echoing the sentiments expressed by HCWs. Many policymakers recognized the pivotal role of HCWs in educating patients, asserting that informed healthcare professionals can significantly impact vaccine acceptance. They also highlighted the importance of community leaders and social media in shaping opinions, as one policy maker noted that *“family and friends can create fear”* (Policymaker, KII). To address hesitancy, policymakers recommended that increased awareness campaigns and reliable information dissemination be implemented, stressing the need for ongoing education and training for HCWs to build trust and counteract misconceptions surrounding vaccines, particularly for those living with HIV.

### Differences and similarities among study populations

While all groups recognized the importance of vaccination, perceptions of COVID-19 vaccines varied, and hesitancy surrounding COVID-19 vaccines was more pronounced among pregnant women, largely due to fear of safety during pregnancy, which was fueled by misinformation from family and friends. HCWs played an important role in influencing vaccine uptake, but their mixed perceptions could either facilitate or hinder acceptance. Policymakers stressed the need for stronger public health interventions, emphasizing education and trust-building strategies to enhance vaccine confidence.

There was a general awareness of vaccines across all study populations, with strong knowledge of routine maternal vaccines such as the tetanus toxoid vaccine. Pregnant women recognized the protective benefits of vaccines for both themselves and their infants, while HCWs and policymakers reinforced the importance of vaccination in disease prevention. However, when it came to COVID-19 vaccination, awareness did not always translate into acceptance.

A key point of divergence emerged in the perception of COVID-19 vaccines. HCWs and policymakers largely viewed the vaccine as essential for protecting pregnant women and also acknowledged concerns regarding its safety and long-term effects. Pregnant women, on the other hand, were more skeptical, with many expressing fear about potential adverse effects and uncertainty about the vaccine’s impact on their pregnancy. Some even doubted the existence of COVID-19, further contributing to hesitancy.

HCWs reported that pregnant women typically accepted vaccines administered during ANC visits, trusting their recommendations. However, this trend did not extend to the COVID-19 vaccine, which was met with widespread hesitancy due to misinformation, fear of side effects, and concerns about its rapid development. Policymakers and HCWs acknowledged that misinformation, particularly from family, friends, and social media, played a significant role in shaping public perceptions and limiting vaccine uptake.

HCWs were identified as key influencers in vaccine decision-making, as pregnant women generally relied on their guidance for ANC-related vaccinations. Policymakers also emphasized the importance of HCWs in educating patients and reducing misinformation. However, some HCWs themselves have doubts about the COVID-19 vaccine’s efficacy, which could further contribute to public hesitancy. Table 1 documents the narratives and relevant quotes about knowledge and perception of COVID-19 vaccination.

**Table 1:**
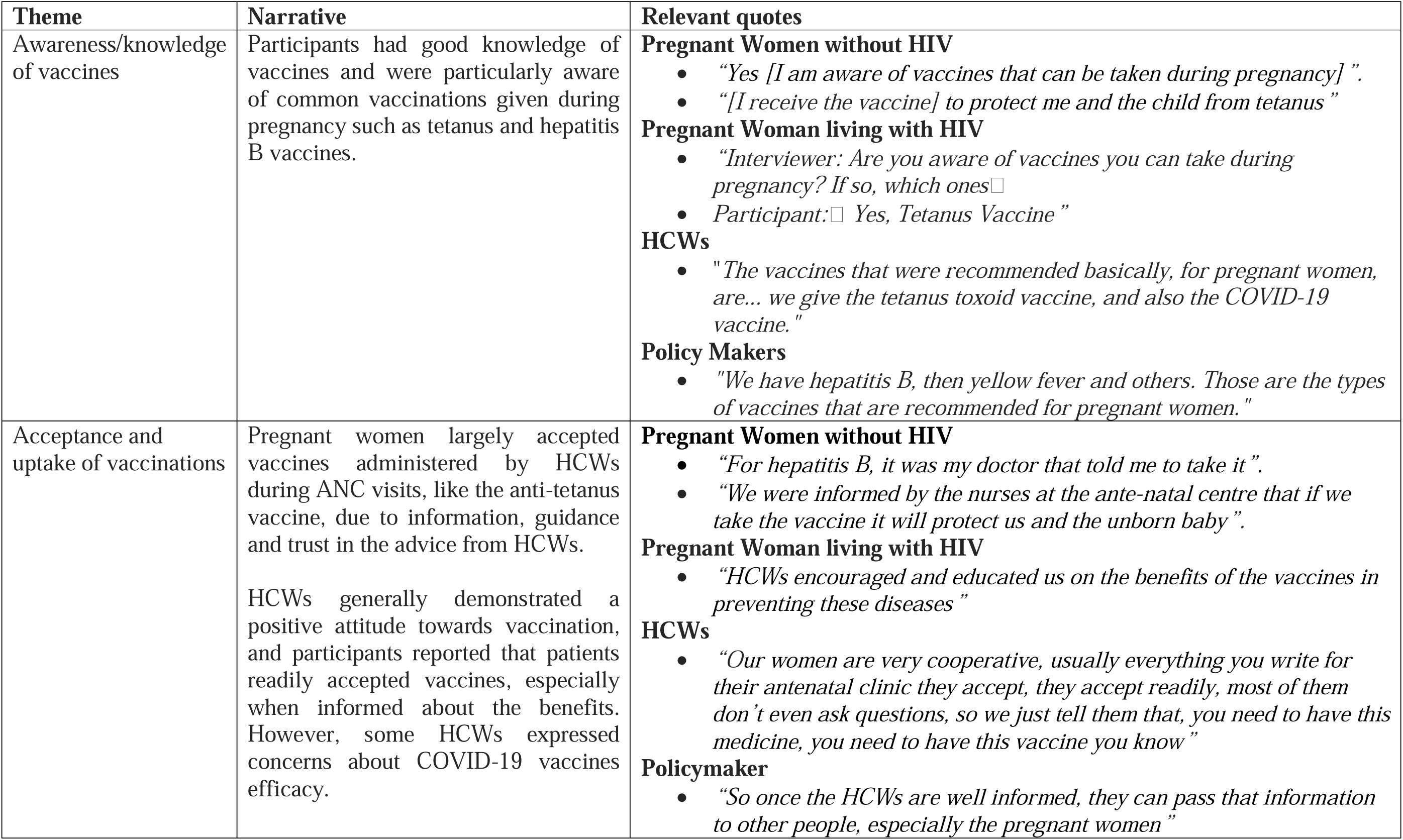
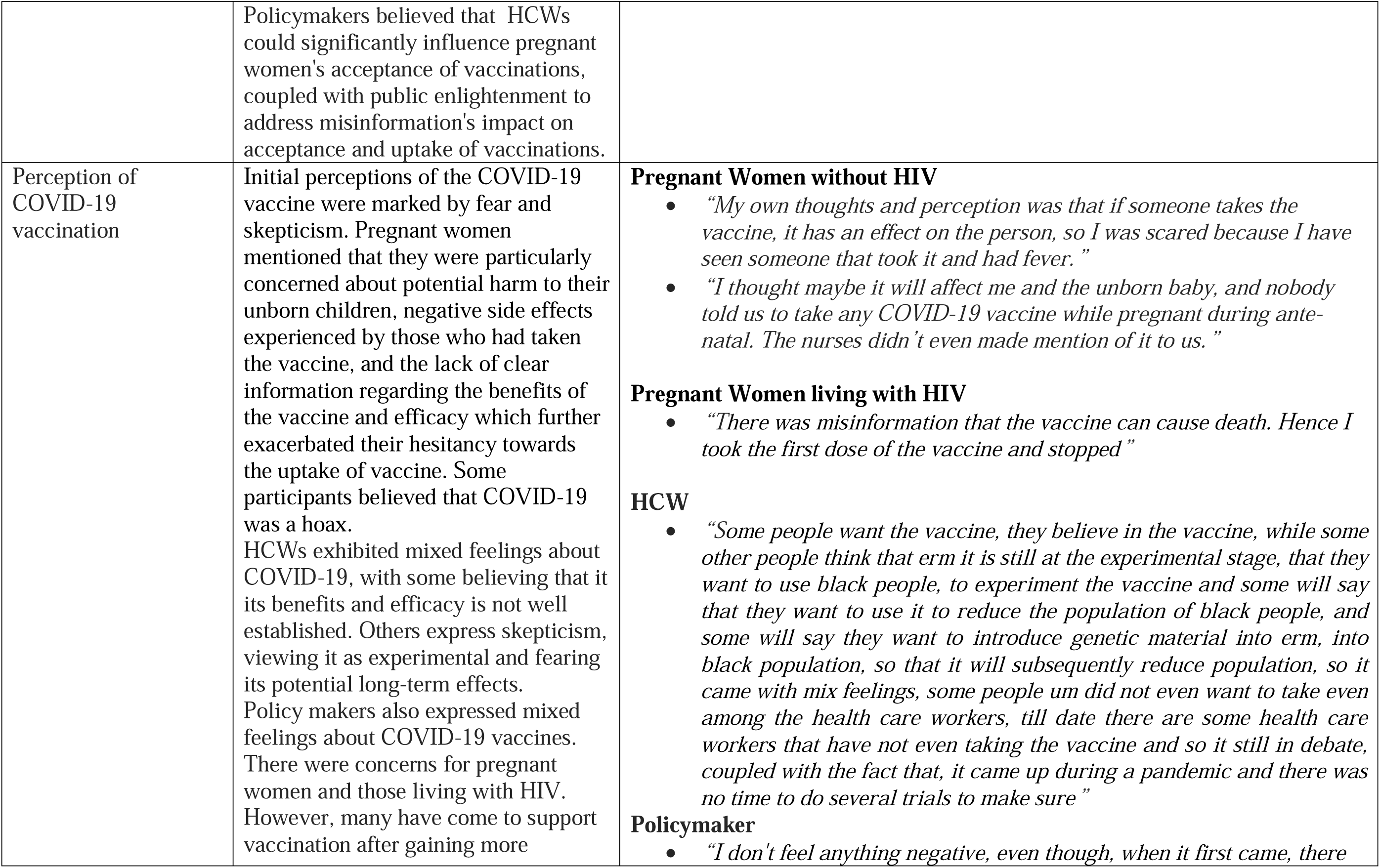
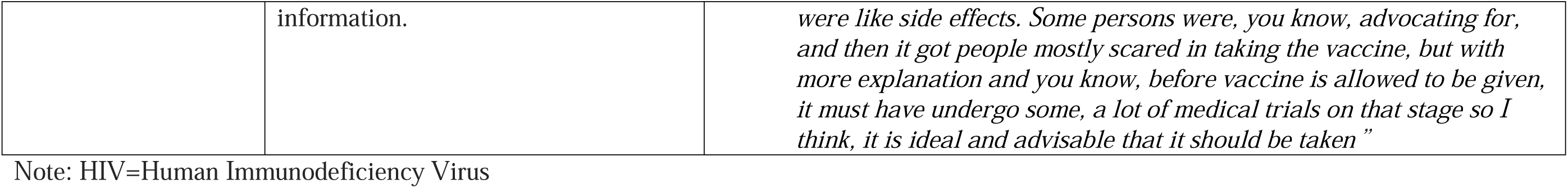
Summary narrative on vaccine knowledge and perception towards COVID-19 vaccination.

| Theme | Narrative | Relevant quotes |
| --- | --- | --- |
| Awareness/knowledge of vaccines | Participants had good knowledge of vaccines and were particularly aware of common vaccinations given during pregnancy such as tetanus and hepatitis B vaccines. | **Pregnant Women without HIV**<br>• “Yes [I am aware of vaccines that can be taken during pregnancy]”.<br>• “[I receive the vaccine] to protect me and the child from tetanus”<br>**Pregnant Woman living with HIV**<br>• “Interviewer: Are you aware of vaccines you can take during pregnancy? If so, which ones □<br>• Participant: □ Yes, Tetanus Vaccine”<br>**HCWs**<br>• “The vaccines that were recommended basically, for pregnant women, are... we give the tetanus toxoid vaccine, and also the COVID-19 vaccine.”<br>**Policy Makers**<br>• “We have hepatitis B, then yellow fever and others. Those are the types of vaccines that are recommended for pregnant women.” |
| Acceptance and uptake of vaccinations | Pregnant women largely accepted vaccines administered by HCWs during ANC visits, like the anti-tetanus vaccine, due to information, guidance and trust in the advice from HCWs.<br>HCWs generally demonstrated a positive attitude towards vaccination, and participants reported that patients readily accepted vaccines, especially when informed about the benefits. However, some HCWs expressed concerns about COVID-19 vaccines efficacy. | **Pregnant Women without HIV**<br>• “For hepatitis B, it was my doctor that told me to take it”.<br>• “We were informed by the nurses at the ante-natal centre that if we take the vaccine it will protect us and the unborn baby”.<br>**Pregnant Woman living with HIV**<br>• “HCWs encouraged and educated us on the benefits of the vaccines in preventing these diseases”<br>**HCWs**<br>• “Our women are very cooperative, usually everything you write for their antenatal clinic they accept, they accept readily, most of them don’t even ask questions, so we just tell them that, you need to have this medicine, you need to have this vaccine you know”<br>**Policymaker**<br>• “So once the HCWs are well informed, they can pass that information to other people, especially the pregnant women” |
|  | Policymakers believed that HCWs could significantly influence pregnant women's acceptance of vaccinations, coupled with public enlightenment to address misinformation's impact on acceptance and uptake of vaccinations. |  |
| Perception of COVID-19 vaccination | Initial perceptions of the COVID-19 vaccine were marked by fear and skepticism. Pregnant women mentioned that they were particularly concerned about potential harm to their unborn children, negative side effects experienced by those who had taken the vaccine, and the lack of clear information regarding the benefits of the vaccine and efficacy which further exacerbated their hesitancy towards the uptake of vaccine. Some participants believed that COVID-19 was a hoax.<br>HCWs exhibited mixed feelings about COVID-19, with some believing that its benefits and efficacy is not well established. Others express skepticism, viewing it as experimental and fearing its potential long-term effects.<br>Policy makers also expressed mixed feelings about COVID-19 vaccines. There were concerns for pregnant women and those living with HIV. However, many have come to support vaccination after gaining more | **Pregnant Women without HIV**<br>• *“My own thoughts and perception was that if someone takes the vaccine, it has an effect on the person, so I was scared because I have seen someone that took it and had fever.”*<br>• *“I thought maybe it will affect me and the unborn baby, and nobody told us to take any COVID-19 vaccine while pregnant during ante-natal. The nurses didn’t even made mention of it to us.”*<br>**Pregnant Women living with HIV**<br>• *“There was misinformation that the vaccine can cause death. Hence I took the first dose of the vaccine and stopped”*<br>**HCW**<br>• *“Some people want the vaccine, they believe in the vaccine, while some other people think that erm it is still at the experimental stage, that they want to use black people, to experiment the vaccine and some will say that they want to use it to reduce the population of black people, and some will say they want to introduce genetic material into erm, into black population, so that it will subsequently reduce population, so it came with mix feelings, some people um did not even want to take even among the health care workers, till date there are some health care workers that have not even taking the vaccine and so it still in debate, coupled with the fact that, it came up during a pandemic and there was no time to do several trials to make sure”*<br>**Policymaker**<br>• *“I don't feel anything negative, even though, when it first came, there* |
|  | information. | *were like side effects. Some persons were, you know, advocating for, and then it got people mostly scared in taking the vaccine, but with more explanation and you know, before vaccine is allowed to be given, it must have undergo some, a lot of medical trials on that stage so I think, it is ideal and advisable that it should be taken”* |

Note: HIV=Human Immunodeficiency Virus

### Factors that Influence Vaccine Acceptance and Uptake

#### Trust in Healthcare Workers

Trust in HCWs was a crucial factor in vaccine decision-making. Pregnant women generally accepted medications, including vaccines, recommended by HCWs. For HIV-positive women, advice from HCWs whom they trusted influenced decisions to get vaccinated, particularly when information was given about potential benefits and efficacy. However, anti-vaccine sentiments within the hospital settings were also barriers to pregnant women receiving vaccines. A pregnant woman who worked as an admin staff at a different hospital remarked “*I work in a health facility; some persons did not take the vaccine. I collected one dose and did not complete the dose because of their talk against the vaccine*” (Pregnant woman, FGD).

#### Family and friends

Participants’ comments indicated that family and friends influenced vaccine acceptance and uptake among pregnant women, particularly regarding the COVID-19 vaccine. While some participants were actively encouraged to get vaccinated, others were discouraged by the negative experiences of their family members, such as the adverse side effects from the vaccine, leading to hesitancy. One participant expressed fear after hearing about her husband’s severe reaction to the vaccine.

#### Socio-cultural and religious influences

Cultural and religious beliefs were not considered significant barriers to vaccine uptake, especially for pregnant participants. However, personal opinions and misinformation were more influential in their choices to receive the vaccine. A pregnant participant remarked, *“My culture did not stop me from taking any vaccine, but I think it is a personal belief not to take a vaccine”* (Pregnant women, FGD). However, participants suggested that the involvement of religious and community leaders in promoting vaccines is a good strategy to increase acceptance and uptake. Table 2 shows narratives and quotes from the study participants.

**Table 2:**
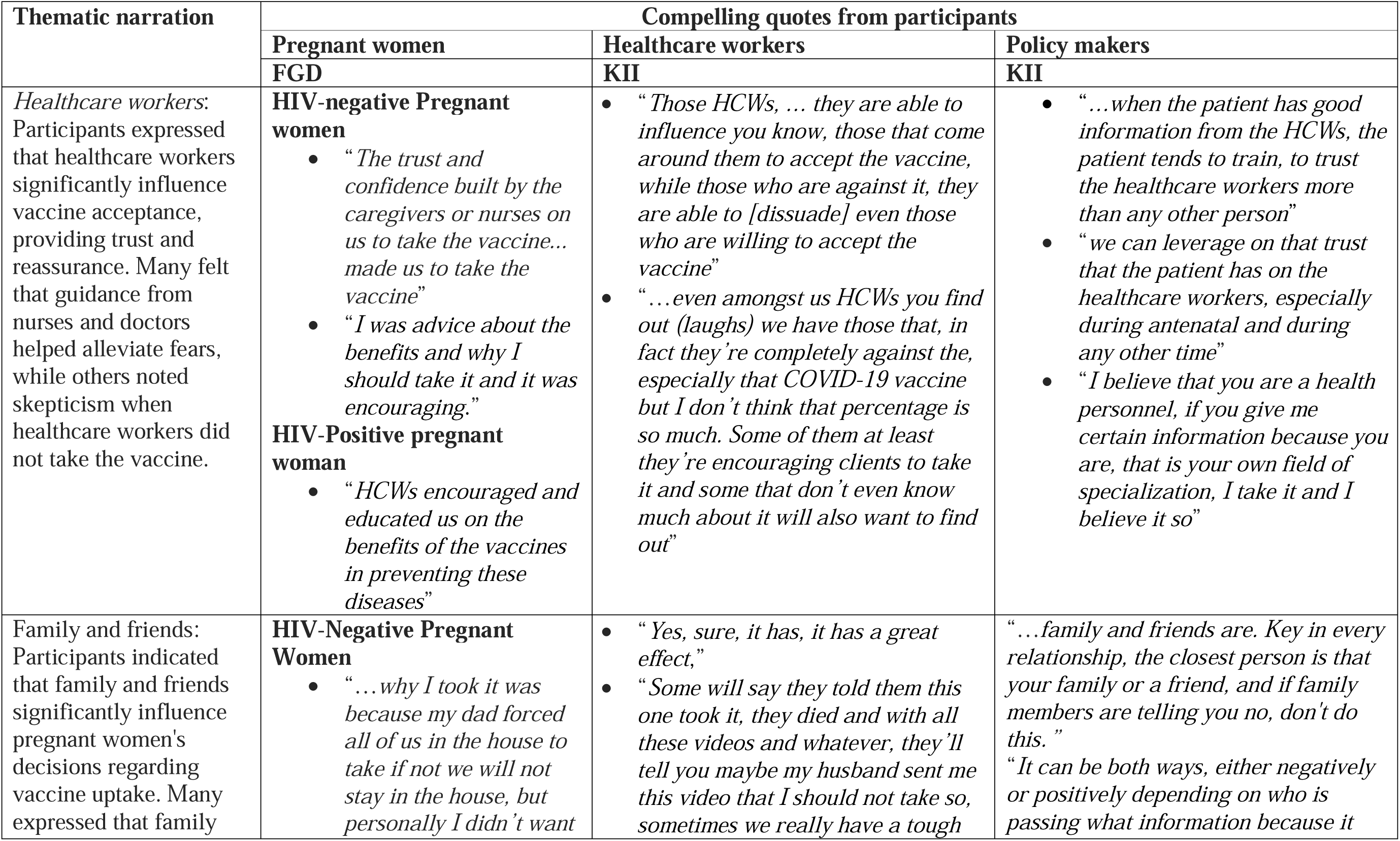
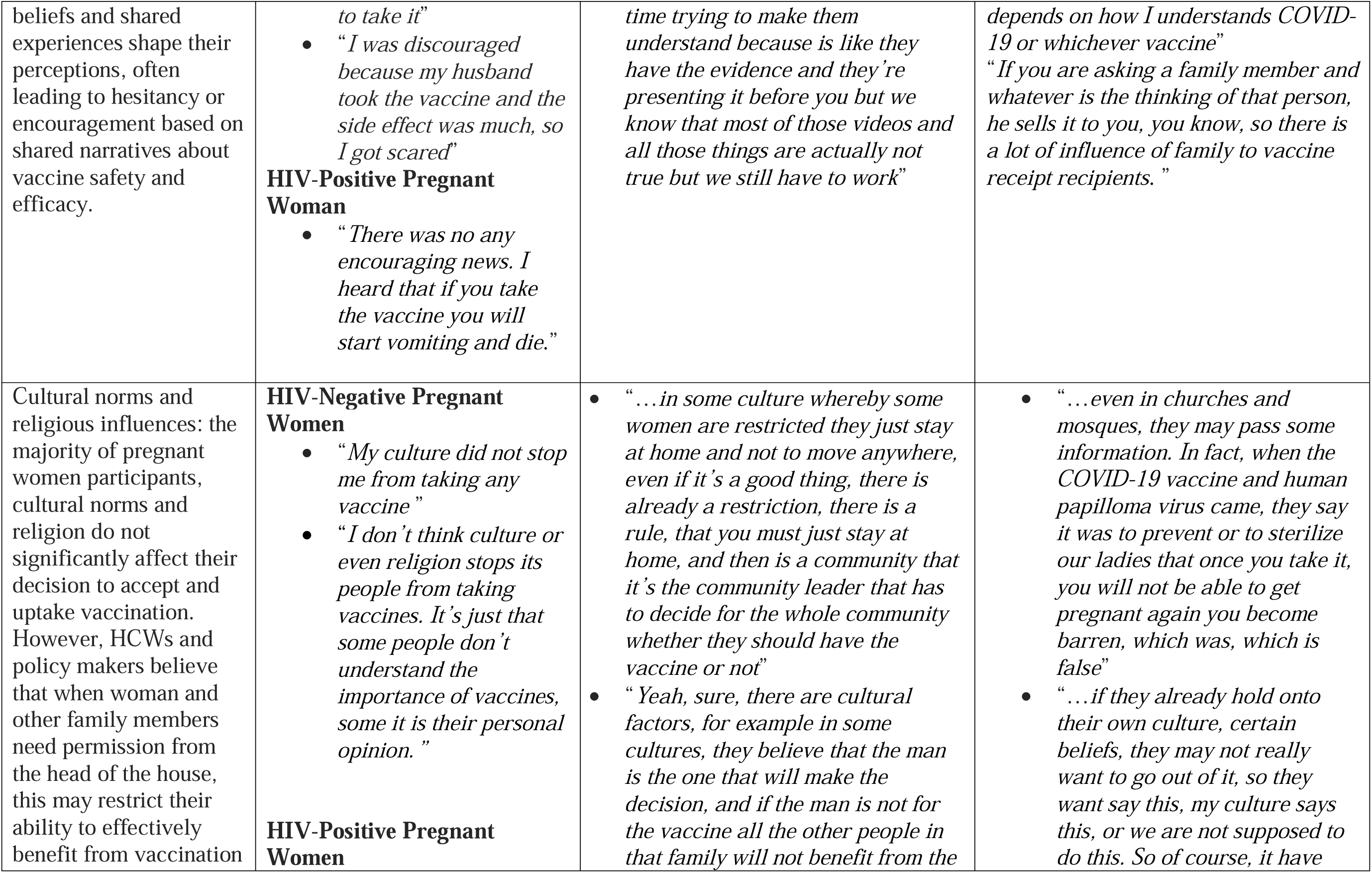
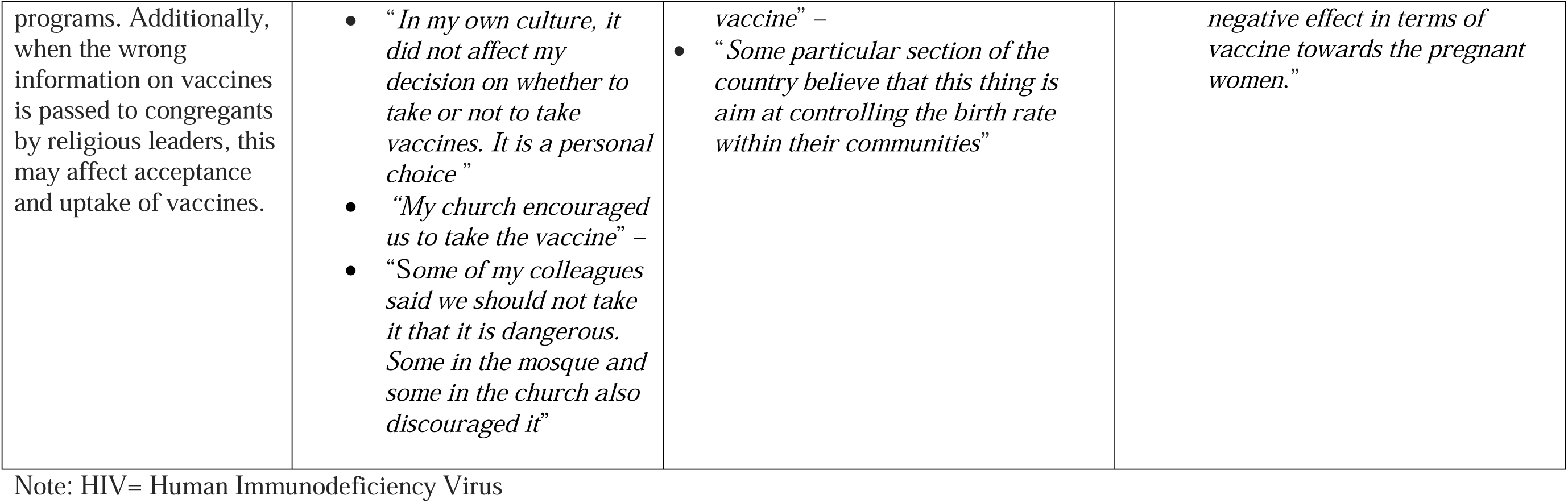
Summary narrative on factors impacting vaccine acceptance and uptake.

## DISCUSSION

This study investigated the knowledge, perceptions of, and acceptance of COVID-19 vaccines among pregnant women, HCWs, and policymakers. The study indicated that pregnant women demonstrated good knowledge of vaccines, particularly tetanus vaccine, with the opinion that this vaccine can provide protection for themselves and their unborn children. This aligns with previous studies conducted in Nigeria, where awareness of routine vaccines, such as tetanus and hepatitis B, was generally high among pregnant women (12–14). However, while knowledge of vaccines was present, the acceptance of the COVID-19 vaccine was marred by skepticism and misinformation.

While the majority of the pregnant women had good knowledge of and acceptance of vaccines for other diseases like tetanus and understood, their role in protecting mothers and children from infection, COVID-19 was faced with negative perceptions that were characterized by the feeling of fear of vaccine safety, adverse events, and misinformation. This finding reinforced the results from a systematic review of 43 studies on pregnant women that reported concerns about vaccine safety and potential harm to their fetuses, which contributed to vaccine hesitancy (15). The finding also supported another study in Pakistan that reported prevalence of vaccine hesitancy amongst pregnant women, which is most significantly affected by the perceptions about the vaccine efficacy, concern about the protection of the fetus and fear of hospitalization due to COVID-19 vaccine intake (16). In addition, our finding is also supported by studies in Congo and Australia that suggest that the the major drivers of vaccine hesitancy were perceived risk to pregnant women and the fetus, poor availability of health information, and inconvenience in receiving vaccines (17),(18). Vaccine hesitancy amongst pregnant women has maternal and child health implications as it increases the risk of severe COVID-19 disease and undermines the public health approach to control the pandemic.

HCWs play a very significant role in addressing COVID-19 vaccine hesitancy among pregnant women. In this study, HCWs demonstrated adequate knowledge of vaccines and had awareness about vaccination for pregnant women. This is consistent with a study in Nigeria, which found that 77% of the HCWs had good knowledge about COVID-19 vaccines (19). However, HCWs were skeptical about COVID-19 vaccines in terms of efficacy, safety, and available information, which can negatively affect the motivation of HCWs to encourage the uptake of COVID-19 vaccines during pregnancy. However, HCWs were reported that pregnant women were compliant with their recommendations for medications without resistance. This finding is similar to a study in Spain where pregnant women reported that HCWs recommendation facilitated their choices of pregnant women to take COVID-19 vaccines (20). Policymakers in our study also reported the significant role HCWs play in encouraging vaccine acceptance, while it is important to ensure safety and efficacy. Thus, it is important that HCWs are adequately trained, given their role in communicating health information to this high-risk population.

The perceptions of COVID-19 vaccination among pregnant women were influenced by fears regarding safety and efficacy. Many participants expressed concerns about potential adverse effects on their health and that of their unborn children. This is similar to findings in a study in Port-Harcourt Nigeria, where the majority of pregnant women expressed safety concerns for the mother and unborn baby as the main reason for non-acceptance of the COVID-19 vaccine (21). This skepticism has also been identified in other studies, where vaccine hesitancy is linked to increased public concerns about vaccine efficacy, availability, and safety, as well as fears surrounding new vaccine technologies and rapid development timelines (9,22).

In our study, HCWs influenced the choices of pregnant women to take COVID-19 vaccines. However, unvaccinated people and anti-vaccine sentiments around the hospital setting contributed to vaccine hesitancy. The belief among some participants that COVID-19 was a hoax further complicated the landscape of vaccine acceptance. This aligns with findings from other studies that indicate a correlation between conspiracy beliefs and vaccine hesitancy (10). The mixed feelings expressed by pregnant women and some HCWs regarding the COVID-19 vaccine also resonate with findings from various studies, where healthcare providers exhibited both support for vaccination and apprehension about its long-term effects (23,24). This study showed that while pregnant women generally accepted routine vaccinations during antenatal care, the uptake of the COVID-19 vaccine was lower. This reflects a broader trend observed during the pandemic, where despite high levels of knowledge, there was a lag in vaccination rates due to fears and misinformation (11,25). The trust in HCWs, however, played a pivotal role in vaccine acceptance, as many women relied on the guidance provided by HCWs. This finding is consistent with the literature, which emphasizes the importance of healthcare provider recommendations in influencing patient decisions regarding vaccinations (26,27).

The influence of family and friends was identified as a significant factor impacting vaccine acceptance. This aligns with social norms theory, which posits that individuals are heavily influenced by the beliefs and behaviors of those around them (28). In this context, negative narratives about the COVID-19 vaccine shared by family members contributed to non-acceptance, a phenomenon also documented in other research (29,30). Also, other factors, such as trust in HCWs and misinformation from family and friends, influenced COVID-19 hesitancy among pregnant women. The role of HCWs as trusted sources of information is well-documented, as studies have shown that patients are more likely to accept vaccinations when they perceive their HCWs as credible and knowledgeable (24,31,32). Conversely, misinformation remains a significant barrier to vaccine uptake as many participants were influenced by false narratives regarding the COVID-19 vaccine, including beliefs that it was experimental, harmful or intended for population control. This is consistent with findings from the World Health Organization, which reported that misinformation significantly hindered vaccination efforts globally (7).

This study has some limitations. We adopted a qualitative design, utilizing a small sample size. As a result, findings cannot be generalized to the entire population of pregnant women, HCWs and policymakers in Nigeria. However, this study provides in-depth insights into participant’s perceptions and can be used to drive public health policies to address vaccine hesitancy amongst pregnant women and provides useful insights for subsequent vaccination programs.

## CONCLUSION

The findings on vaccine knowledge, perception, and acceptance among pregnant women, HCWs, and policymakers in Nigeria reveal a complex interaction of factors influencing vaccination behaviors. While knowledge of vaccines is generally high, skepticism about the COVID-19 vaccine remains a significant barrier to uptake. Trust in healthcare providers, the influence of family and friends, and the prevalence of misinformation are critical factors shaping vaccine acceptance.

To improve COVID-19 vaccine acceptance and vaccination rates, it is pertinent to address misinformation through targeted public health education campaigns and to leverage the trust that pregnant women place in HCWs. Additionally, active and ongoing engagement of community and religious leaders in vaccination advocacies and exercises could further enhance vaccine acceptance and uptake. Continued research is needed to explore the evolving perceptions of vaccines in the context of emerging health crises.

## Data Availability

All data produced in the present study are available upon reasonable request to the authors

